# Apnea and hypopnea event detection using EEG, EMG, and sleep stage labels in a cohort of patients with suspected sleep apnea

**DOI:** 10.1101/2024.10.24.24316077

**Authors:** Danny H. Zhang, Jeffrey Zhou, Joseph D. Wickens, Andrew G. Veale, Luke E. Hallum

## Abstract

Automating the screening, diagnosis, and monitoring of sleep apnea (SA) is potentially clinically useful. We present machine-learning models which detect SA and hypopnea events from the overnight electroencephalogram (EEG) and electromyogram (EMG), and we explain detection mechanisms. We tested four models using a novel data set comprising six-channel EEG and two-channel EMG recorded from 26 consecutive patients; recordings were expertly labeled with sleep stage and apnea/hypopnea events. For Model 1, EEG subband power and sample entropy were features used to train and test a random forest classifier. Model 2 was identical to Model 1, but we used EMG, not EEG. Model 3 was a simple decision strategy contingent upon sleep stage label. Model 4 was identical to Model 1, but we used EEG subband power, sample entropy, and sleep stage label. All models performed above chance (Matthews correlation coefficient, MCC > 0): Model 4 (leave-one-patient-out cross-validated MCC = 0.314) outperformed Model 3 (0.230) which outperformed Models 2 and 1 (0.147 and 0.154, respectively). Results indicate that sleep stage label alone is sufficient to detect apnea/hypopnea events. Either EMG or EEG subband power and sample entropy can be used to detect apnea/hypopnea events, but these EEG features likely reflect contamination by EMG. Indeed, EMG power was modulated by apnea/hypopnea event beginning and end, and similar modulation appeared in EEG power. Machine-learning approaches to the detection of apnea/hypopnea events using overnight EEG must be explainable; they must account for EMG contamination and sleep stage.

## 1 Introduction

Sleep apnea (SA) is a serious breathing disorder involving repeated apneic and hypopneic events during sleep. SA is common; in the United States, at least mild forms of the disorder affect more than 17.4% of women and 33.9% of men between 30 and 70 years of age (Peppard et al., 2013; Gottlieb and Punjabi, 2020). Sufferers of SA are at increased risk of workplace and motor vehicle accidents (Ulfberg et al., 2000), and SA is associated with a range of serious, chronic diseases, including cardiovascular disease and diabetes (Jennum & Riha, 2009; Pumjabi, 2008; Sullivan et al., 1981; Marin et al., 2005). Continuous positive airway pressure (CPAP) — the gold standard SA treatment — is highly effective (Sullivan et al., 1981), however, there are barriers to treatment. SA is diagnosed using polysomnography (PSG) manually annotated by expert sleep physiologists (Rundo et al., 2019). PSG is instrumentation-intensive and therefore, may interfere with standard patterns of sleep (Park & Choi, 2019). Furthermore, demand for overnight PSG exceeds supply; wait times for those requiring screening and diagnosis can range from 2 to 60 months (Flemmons et al., 2004), and it is estimated that a large proportion of SA sufferers are undiagnosed (Young et al., 1997).

Tools to aid detection, diagnosis, and treatment of SA are, therefore, a worthwhile pursuit, especially those involving simplified instrumentation and automation, with potential for use in the home as well as the clinic (Mykytyn et al., 1999; Brunyneel et al., 2011; Mikkelsen et al., 2017). Automated systems making use of explainable machine-learning (Holzinger, 2016) could, potentially, be used to expedite and augment diagnostic and treatment decisions; explainable systems are those wherein mechanisms of detection are made available to clinicians. Furthermore, understanding the electroencephalographic correlates of SA may be of general benefit to sleep medicine; potentially, EEG markers may be used to phenotype patient groups, thus indicating disease states and predicting treatment outcomes (D’Rozario et al., 2017).

Several studies have reported above-chance detection of apnea/hypopnea events using overnight EEG recordings (Almuhammadi et al., 2015; Bello & Alqasemi, 2021; Barnes et al., 2022; Prucnal & Polak, 2019a; Prucnal & Polak, 2019b; Wang et al., 2020; Zhao et al., 2021). Noteworthy amongst these studies is recent work by Zhao and colleagues (2021). They analysed two-channel EEG recordings from 30 SA patients, computing subband power and sample entropy, and used these computed features to perform ternary classification (i.e., to classify epochs as reflecting events of (1) obstructive SA, (2) central SA, and (3) normal breathing). Using a cross-validation framework that pooled data across patients, they showed that a random forest (RF) classifier (accuracy equal to 89.0%) outperformed both a K-nearest neighbor classifier and a support-vector machine.

The above-cited work by Zhao and colleagues (2021), as well as similar studies, lacks mechanistic explanation. This is problematic, because it remains unclear whether there exists an EEG marker specific to apnea/hypopnea events which is sufficiently robust to enable classification that generalises to previously unseen patients (“patient-wise” classification). Because, in general, the frequency of apnea/hypopnea events varies as a function of sleep stage (Cartwright et al., 1991; Ratnavadivel et al. 2009), it is possible that the EEG signals enabling the above-cited classification may not be EEG markers specific to apnea/hypopnea events, but rather classification may depend on EEG signals characteristic of the different sleep stages (the electroencephalographic correlates of sleep stage are reviewed by Carskadon & Dement, 2017). Furthermore, because, in general, electromyographic (EMG) signals are known to contaminate EEG (Goncharova et al., 2003), it is possible that the putative EEG signals enabling the above-cited classification are not, in fact, EEG signals (a point we take up in the Discussion). Despite this lack of mechanistic explanation, it is nonetheless reasonable to suppose that apnea/hypopnea events can be detected using overnight EEG. Using power spectral analysis of EEG recordings, several groups have reported robust differences between cohorts of SA sufferers and normal controls, or between cohorts of sufferers of mild and more severe disease (reviewed by D’Rozario et al., 2017; reviewed by Puskás et al., 2017). Furthermore, the termination of events of apnea/hypopnea is known to be associated with arousal — often cortical arousal reflected in EEG (Remmers et al., 1978; reviewed by Eckert & Younes, 2014; reviewed by Deegan and McNichols, 1995).

Here, we are interested in developing a mechanistic understanding of the putative EEG signals that enable detection of apnea/hypopnea events. We tested two hypotheses:

1. We hypothesized that sleep stage is sufficient to detect epochs containing events of apnea/hypopnea. This hypothesis is plausible: apnea/hypopnea events are not uniformly distributed across stage, and therefore information about sleep stage carries information about apnea/hypopnea.
2. We hypothesized that EMG (not EEG) subband power and sample entropy is sufficient to detect epochs containing apnea/hypopnea events. This hypothesis is plausible: EMG is known to contaminate EEG.

Below, we show that both these hypotheses pass the test. This indicates that neither EEG subband power nor sample entropy are markers specific to SA enabling robust classification that generalises to previously unseen patients. Machine learning using overnight EEG for apnea/hypopnea event detection must be explainable; it must account for EMG contamination and sleep stage.

## 2 Methodology

### 2.1 EEG and EMG recordings

EEG and EMG recordings were taken from overnight polysomnograms (PSGs) acquired at the New Zealand Respiratory & Sleep Institute (NZRSI) and annotated by experienced sleep physiologists. The patients we report here are consecutive in that the data set was analyzed in the order it was provided to the principal investigator (LH) by sleep physiologists who were naive to the study’s hypotheses. EEG recordings were made using a standard, six-channel, polysomnographic montage referenced to the contralateral mastoids: O1, O2, C3, C4, F3, F4. EMG recordings were made using a standard, two-channel, polysomnographic montage similarly referenced: E1, E2. EEG and EMG signals were recorded at 200 samples per second. Because this study involved the retrospective analysis of clinical EEG and EMG previously acquired at NZRSI and provided to the principal investigator (LH) in de-identified format, the Auckland Health Research Ethics Committee waived the requirement for any further approval to use these datasets in this study.

### 2.2 Feature extraction

These EEG and EMG recordings had been segmented by sleep physiologists into 30-second-long contiguous epochs; each epoch was therefore labelled with one of five sleep stages: wake, REM, N1, N2, N3 (Figure 1). For each epoch, we computed a vector comprising 10 features as follows. First, we used a robust method to standardise each electrophysiological recording: we subtracted the median, and then divided by 0.7413-times the interquartile range (“robust standardisation”). Second, we filtered each epoch to produce five subband signals. The filters were 3rd-order Butterworth filters, with nominal and actual cut-off frequencies tabulated in Table 1. Third, for each subband signal, we calculated the variance (i.e., the power) and the sample entropy (Richman & Moorman, 2000). To calculate the sample entropy, we used an implementation provided by Martínez-Cagigal (2018), and parameters identical to those of Zhao et al. (2021). Thus, each 30-second epoch on each recording channel was reduced to a vector comprising 10 features: power and sample entropy in each of five subbands. We did the filtering and computed the features using Matlab (MathWorks Inc., Natick, MA, United States; version 9.13.0.2126072 (R2022b) Update 3).

**Table 1:** Third-order Butterworth filters used to isolate subband signals.

|  | Delta | Theta | Alpha | Beta | Gamma |
| --- | --- | --- | --- | --- | --- |
| Nominal frequency range (Hz) | < 4 | 4 to 8 | 8 to 13 | 13 to 30 | > 30 |
| Actual frequency range (Hz; 6dB down) | < 4.8 | 3.7 to 8.5 | 7.6 to 13.6 | 12.0 to 32.1 | > 28.5 |

Abbreviations: Hz, Hertz; dB, decibels

**Figure 1:**
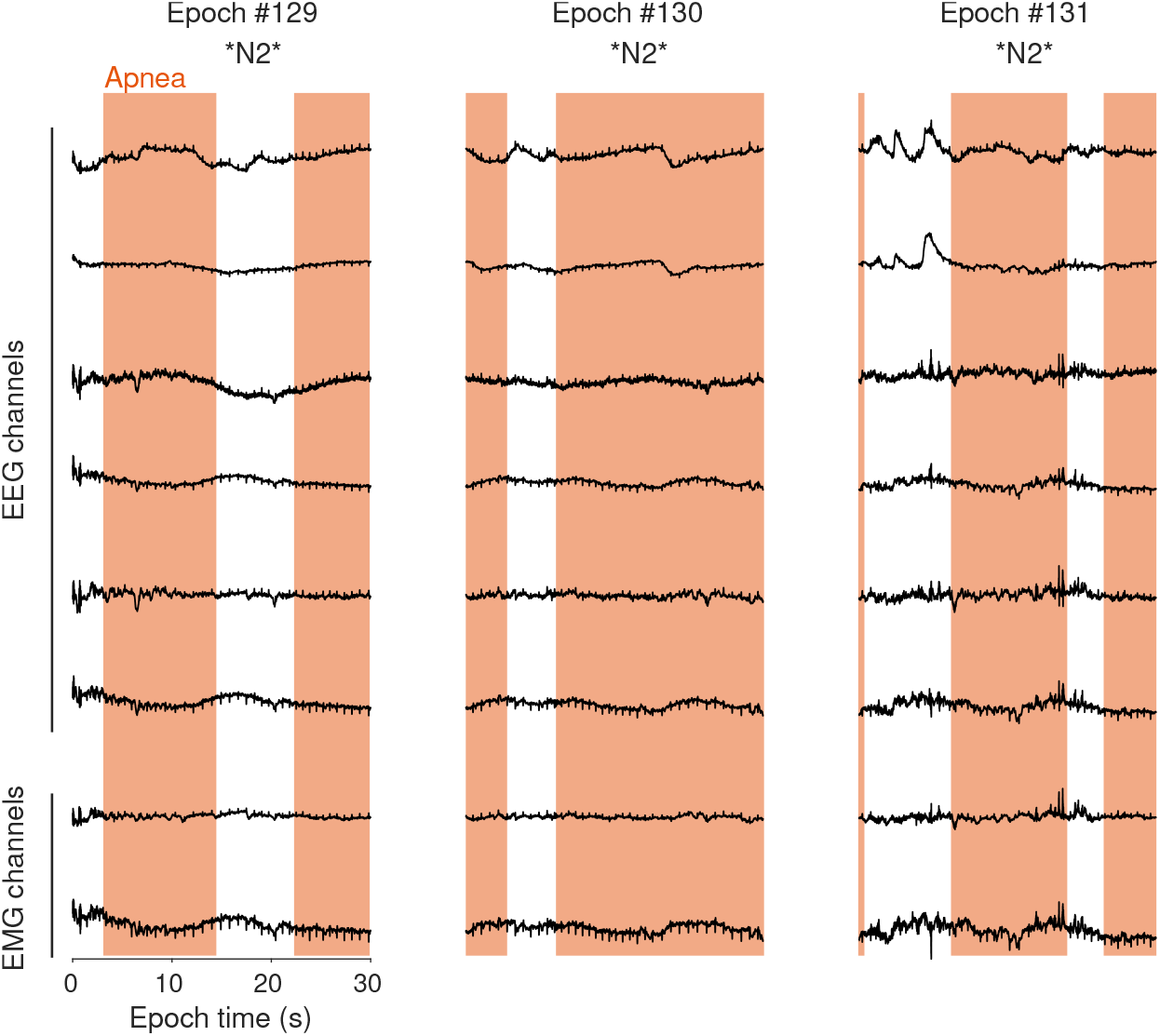
Example electroencephalogram (EEG) and electromyogram (EMG) recordings drawn from patient P1’s polysomnogram (PSG). These recordings are raw and unfiltered. Rows show different recording channels (upper six: EEG; lower two: EMG), and columns show three temporally adjacent, 30-second epochs (Epoch #129, #130, and #131). These epochs were recorded approximately one hour into the patient’s overnight sleep episode. Using the entire PSG, an expert physiologist labeled each epoch with sleep stage (three labels, “N2”, enclosed by asterisks) and demarcated apnea/hypopnea events (five orange intervals). We pre-processed and filtered recordings. For each epoch we then computed a 10-dimensional feature vector (see *Feature extraction*), and we labelled each epoch as belonging to one of two classes: “apnea/hypopnea” or “non-apnea/hypopnea” (see *Classification and cross-validation*). Here, all three epochs are labelled “apnea/hypopnea” because each contains one or more apnea/hypopnea event(s) with cumulative duration >= 10 seconds. We used feature vectors and labels to train and test random forest classifiers (see text).

### 2.3 Classification and cross-validation

To gain mechanistic insight into the detection of apnea/hypopnea events from overnight EEG, we tested four models:

- For **Model 1**, we used EEG subband power and sample entropy (see Feature extraction) to train and test a random forest (RF) classifier. This classifier was composed of 500 decision trees, unpruned, and used 10 predictors (i.e., features) at each split. We implemented this RF classifier using R (version 4.3.2 (2023-10-31); R Core Team, 2023), and made specific use of packages “tree” (version 1.0-43; Ripley, 2023) and “randomForest” (version 4.7-1.1; Liaw & Wiener, 2002). For this model, the number of features used to represent each epoch was 6 channels × 5 subbands × 2 measures (power, sample entropy) = 60.
- **Model 2** was identical to Model 1, except we used EMG (not EEG) subband power and sample entropy to train and test this classifier. For this model, the number of features used to represent each epoch was 2 channels × 5 subbands × 2 measures (power, sample entropy) = 20.
- **Model 3** was a simple decision strategy contingent on sleep stage label. There were 5 sleep stage labels: Wake, REM, N1, N2, N3.
- **Model 4** was identical to Model 1, except we used EEG subband power and sample entropy, as well as sleep stage label as features to train and test this classifier. To represent sleep stage label as a feature, we used a length-five one-zero vector wherein each component represented a stage. For example, sleep stage REM was represented as [0 1 0 0 0], whereas sleep stage N1 was represented as [0 0 1 0 0]. Therefore, for this model, the number of features used to represent each epoch was 60 from Model 1 + 5 = 65.

We used intervals of apnea/hypopnea expertly demarcated by sleep physiologists to label each epoch as belonging to one of two classes: epochs containing one or more apnea/hypopnea event(s) with cumulative duration >= 10 seconds were labelled “apnea/hypopnea”; other epochs were labelled “non-apnea/hypopnea”. We used leave-one-patient-out cross-validation (i.e., “patient-wise” cross-validation) to quantify the performance of our RF classifier. To do so, on fold one, we used recordings from all but the first patient to train our classifier, and we then tested that trained classifier using recordings from the first patient and recorded its performance. On fold two, we used recordings from all but the second patient to train our classifier, and we then tested that trained classifier using recordings from the second patient and recorded its performance. And so on for folds one through N, where N is the number of patients. The procedure is illustrated in Figure 2.

**Figure 2:**
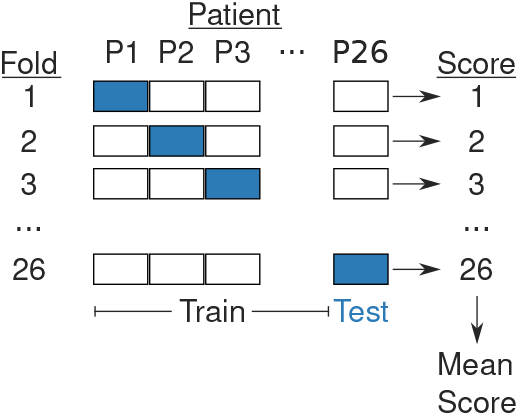
Leave-one-patient-out (“patient-wise”) cross-validation. On each fold of our 26-fold procedure, we trained models using data recorded from 25 patients (white blocks) and tested models using data recorded from the remaining patient (blue). Therefore, our assessment of each model’s performance captured its ability to generalize across patients.

### 2.4 Apnea/hypopnea-triggered broadband instantaneous power

To better understand the association between apnea/hypopnea events and systematic alterations in EEG and EMG recordings, for each patient, we computed an event-triggered average of each recording using the following six-step algorithm:

1. Standardize the EEG (or EMG) recording (see Feature extraction)
2. Segment the standardized recording into *M* 30-second-long epochs (*M* is the number of apnea/hypopnea events experienced by the patient during the sleep episode, and each epoch straddles the start time of an event)
3. For each epoch, subtract the mean and low-pass filter (3rd-order Butterworth filter with cut-off = 48 Hz)
4. For each epoch, square each value (thus giving a measure of instantaneous power) and then square-root each value (to stabilize variance)
5. At each point in time, average across all *M* epochs, giving an event-triggered “average epoch”
6. At each point in time, z-score this average epoch using a null ensemble of 100 average epochs

On step 2 of this algorithm, we segmented each recording such that each resulting epoch straddled the start time of an apnea/hypopnea event, that is, the initial 15 s of each resulting epoch corresponded to the EEG (or EMG) recording before the start of an apnea/hypopnea event, and the subsequent 15 s corresponded to the recording after the start of event. On step 6 of this algorithm, we used a null ensemble of 100 “average epochs”. Each average epoch comprising this null ensemble was computed as described by steps 1 to 5 of the algorithm, but the *M* points in time were randomised (uniformly) across the patient’s sleep episode.

We also computed an event-triggered average using the end times of the *M* apnea/hypopnea events (using an algorithm identical to the above-described “start-time” algorithm).

### 2.5 Classifier performance metrics

To quantify classifier performance, we used MCC, the Matthews correlation coefficient (Matthews, 1975; Chicco and Jurman, 2020; Delgado and Tibau, 2019). We used MCC because Chicco and Jurman (2020) demonstrated its superiority over other classifier performance measures where unbalanced data sets are concerned, that is, data sets wherein the number of observations in one or other class is relatively large. We also used accuracy, sensitivity, and specificity. These latter three metrics are susceptible to bias when data sets are unbalanced; we provide them here primarily for the sake of comparison with other studies. Chicco and Jurman (2020) point out that, when data sets are unbalanced, “accuracy cannot be considered a reliable measure… because it provides an overoptimistic estimation of the classifier ability on the majority class”. It follows that, when data sets are unbalanced, sensitivity (i.e., the true-positive rate) and specificity (i.e., the true-negative rate) are also unreliable.

Metrics were calculated in the standard fashion:

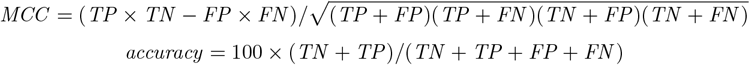

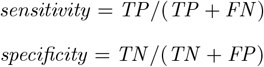

In the above equations, TN, TP, FP, and FN are true negatives, true positives, false positives, and false negatives, respectively.

We used a shuffle test to empirically estimate chance performance. To do so, we shuffled the class labels in the training set; we then re-trained and re-tested the classifier, again using leave-one-patient-out cross-validation. We reasoned that a classifier trained after shuffling labels would be incapable of learning salient EEG or EMG features for apnea/hypopnea event detection, but would nonetheless be capable of learning the statistics of data set imbalance, and capable of biasing its behaviour accordingly.

## 3 Results

We studied overnight EEG and EMG recorded from a cohort of 26 consecutive patients. All patients were enrolled between 8 January and 26 February, 2020. Most patients (n = 22) underwent split-night studies, that is, PSG for the first part of the night (the “analysis period”) during which sleep was measured and SA severity was assessed, followed by CPAP titration for the remaining part of the night. Patients exhibited a range of SA severity: the cohort’s apnoea-hypopnoea index (AHI) ranged from 3.1 to 60.7 events/hour. Therefore, by conventional standards, seven patients suffered mild SA (AHI <= 15), seven patients suffered moderate SA (15 < AHI <= 30), and 12 patients suffered severe SA (AHI > 30). The mean (median) AHI was 29.1 (27.2) events/hour.

Overnight EEG and EMG recordings during the analysis period were segmented into 12,297 30-second epochs. Each of these epochs was labelled by expert sleep physiologists for sleep stage. Sleep physiologists also demarcated periods of apnea and hypopnea during the analysis period. Overall, epochs were most often labelled sleep stage N2 (37.7% of all epochs); epochs were also labelled sleep stage Wake (18.0%), N3 (15.3%), N1 (15.5%), and REM (13.4%). Some 71.7% of all epochs labelled N1 contained one or more apnea/hypopnea event(s) with total duration at least 10 seconds. Stage N1 was the only “majority apnea/hypopnea” sleep stage; for stages N2, Wake, REM, and N3, some 37.9%, 7.9%, 30.2%, and 14.7% of epochs contained one or more apnea/hypopnea event totaling at least 10 seconds, respectively. Data are shown in Figure 3.

**Figure 3:**
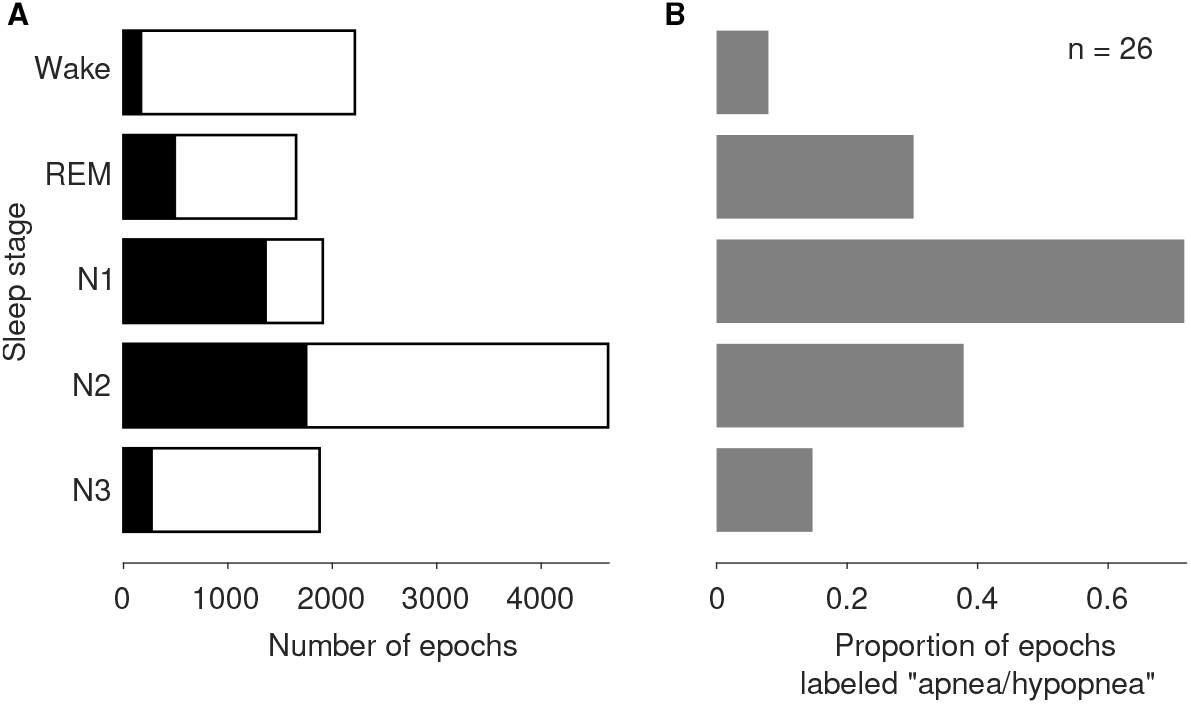
Histograms showing the distribution of sleep stages and apnea/hypopnea events across the cohort (n = 26). (A) The bars represent the 12,297 contiguous 30-second epochs that we analyzed. The filled section (black) of each bar represents epochs containing one or more apnea/hypopnea event(s) with cumulative duration >= 10 seconds. For example, the data set comprised 1,878 epochs labelled N3; some 276 (14.7%) of these epochs contained apnea/hypopnea event(s) with cumulative duration >= 10 seconds. (B) The bars represent the proportion within each sleep stage of epochs that contained apnea/hypopnea event(s) with cumulative duration >= 10 seconds. Stage N1 is the only “majority apnea/hypopnea” sleep stage (i.e., more than 50% of all N1 epochs were labelled “apnea/hypopnea”).

We used EEG subband power and sample entropy (see Feature extraction) to train a random forest (RF) classifier (“Model 1”). We used leave-one-patient-out cross-validation to assess the performance of this model (see Classification and cross-validation). Overall, the Matthews correlation coefficient (MCC) was equal to 0.154 (the mean across folds of our leave-one-patient-out cross-validation), with 95% confidence interval computed via bootstrapping equal to [0.109, 0.214]. In addition, we used EMG subband power and sample entropy to train a second RF classifier (“Model 2”). Overall, the Matthews correlation coefficient (MCC) was equal to 0.147 [0.095, 0.216]. These performance measures are plotted in Figure 4. Other performance metrics, including accuracy, sensitivity, specificity, are tabulated in Table 2.

**Table 2:** Classification models’ performance averaged over our leave-one-patient-out cross-validation procedure (square brackets show the 95% confidence interval of the average immediately above). For MCC, Models 1 and 2 were outperformed by Model 3, which was in turn outperformed by Model 4. MCC is the appropriate performance measure because classes were unbalanced (i.e., in our data set, there were fewer epochs labelled “apnea/hypopnea” than there were epochs labelled “non-apnea/hypopnea”). Bold typeface indicates the best performing model for each metric. The shuffle test (“Control”) is our empirical estimate of chance performance (see text).

|  | Model 1 | Model 2 | Model 3 | Model 4 | Control |
| --- | --- | --- | --- | --- | --- |
| MCC | 0.154<br>[0.109, 0.214] | 0.147<br>[0.095, 0.216] | 0.230<br>[0.181, 0.275] | **0.314**<br>[0.242, 0.410] | -0.024<br>[-0.046, -0.008] |
| Accuracy (%) | 64.986<br>[55.948, 72.721] | 66.512<br>[57.674, 74.063] | 65.537<br>[56.172, 72.648] | **71.134**<br>[62.597, 78.581] | 52.626<br>[39.744, 63.468] |
| Sensitivity | 0.361<br>[0.237, 0.502] | 0.373<br>[0.254, 0.515] | 0.315<br>[0.257, 0.377] | **0.521**<br>[0.380, 0.657] | 0.022<br>[0.007, 0.072] |
| Specificity | 0.799<br>[0.690, 0.886] | 0.774<br>[0.667, 0.855] | 0.916<br>[0.879, 0.942] | 0.820<br>[0.714, 0.891] | **0.967**<br>[0.913, 0.984] |

Abbreviations: MCC, Matthews correlation coefficient

**Figure 4:**
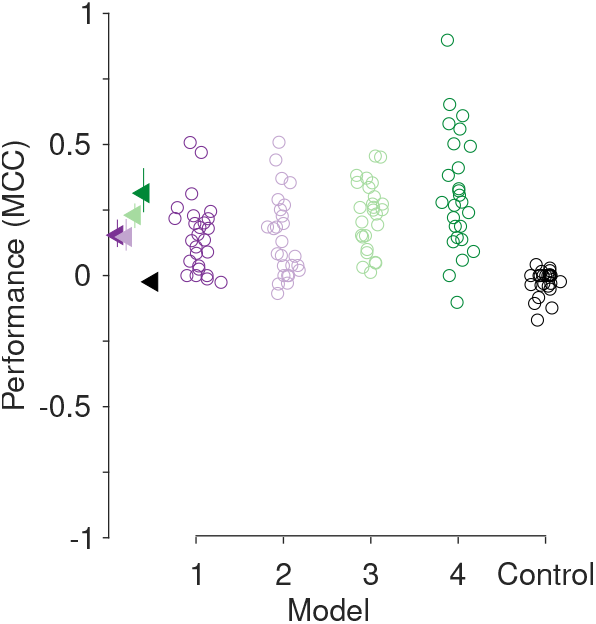
Performance (Matthews correlation coefficient, MCC) of classification models. Our cohort consisted of n = 26 patients; we used leave-one-patient-out cross-validation, and for this reason each model’s performance is represented by 26 symbols: one symbol for each patient. The triangles at left indicate the performance means, and where visible the error bars mark 95% confidence intervals computed with bootstrapping (Efron and Tibshirani, 1994). Overall, Model 4 outperformed Model 3, which outperformed Models 2 and 1. In addition to MCC, other performance metrics are tabulated in Table 2. The shuffle test (“Control”) is our empirical estimate of chance performance; there, labels (“apnea/hypopnea” and “non-apnea/hypopnea”) on training data were shuffled prior to re-training Model 4 (see text).

For the sake of comparison to Zhao et al. (2021), we pooled EEG subband power and sample entropy across patients and then trained a RF classifier. Here, we did not use leave-one-patient-out cross-validation, but rather we held out from training approximately 20% of observations, chosen at random, and later used these observations to assess model performance. This model’s Matthews correlation coefficient (MCC) was equal to 0.73 and the accuracy was equal to 88.5%. We empirically estimated baseline (i.e., chance) performance; when we shuffled apnea/hypopnea labels on the held-in data before training, MCC and accuracy decreased to 0.0 and 66.7%, respectively. This accuracy (88.5%) is comparable to that of the ternary classifier used by Zhao et al. (89.0%; see our Introduction). However, we emphasise that pooled models like that of Zhao et al. lack external validity (a point we take up in our Discussion).

In our data set, apnea/hypopnea events were not uniformly distributed across sleep stages (Figure 3). Therefore, we wondered whether a simple decision strategy contingent on sleep stage label (“Model 3”) could outperform our RF classifiers which used EEG or EMG subband power and sample entropy as features (“Model 1” and “Model 2”). To answer this question, we again used a leave-one-patient-out cross-validation procedure as follows. For patient P1, we used data from patients P2 to P26 to form an empirical distribution, *D*_2_ − _26_, of the proportion within each sleep stage of epochs that contained apnea/hypopnea event(s) with total duration at least 10 seconds; this distribution was similar, but not identical, to the distribution shown in Figure 3(B). Then, we classified P1’s epochs according to this distribution. Specifically, for *D*_2_ −_26_, sleep stage N1 was the only “majority apnea/hypopnea” stage. Therefore, all of P1’s epochs labelled sleep stage N1 were classified as “apnea/hypopnea”, whereas all other epochs were classified as “non-apnea/hypopnea”. We repeated this procedure for each patient. For Model 3, overall, MCC was equal to 0.230 (the mean across patients), with 95% confidence interval equal to [0.181, 0.275]. This performance measure is plotted in Figure 4, and other performance metrics, including accuracy, sensitivity, and specificity, are tabulated in Table 2.

The performance of Model 1 and Model 3 was not correlated. In other words, when Model 1 performed well predicting the presence or absence of apnea/hypopnea events for a particular patient, that did not indicate that Model 3 would also perform well predicting the presence or absence of apnea/hypopnea events in that same patient. This lack of correlation indicated that the Model 1 and Model 3 features could be combined to further improve classification performance. Therefore, we used EEG subband power, sample entropy, and sleep stage label to train a RF classifier (“Model 4”). Overall, the MCC was equal to 0.314 [0.242, 0.410] (Figure 4; Table 2). Since the performance of Model 1 and Model 2 was correlated, it is unsurprising that Model 4 outperformed all models, 1 through 3. (A variation on Model 4, which used EMG subband power, sample entropy, and sleep stage label performed comparably: MCC equal to 0.288 [0.209, 0.399].)

To better understand the mechanisms enabling the performance of Model 1, 2, and 4, we compared subband power in epochs labeled “apnea/hypopnea” with that of epochs labelled “non-apnea/hypopnea”. To do so, for each patient, we pooled EEG subband power from all frequency bands (delta through gamma; see Methods) to form two samples: “apnea/hypopnea” and “non-apnea/hypopnea”. Then, we used a Wilcoxon rank sum test to test for equal medians of these two samples. We used the z-scored rank sum statistic returned from this test as an index of subband power modulation during apnea/hypopnea (subband power modulation index, SPMI). Therefore, more negative SPMI values indicate a more robust difference of subband power during apnea/hypopnea as compared to non-apnea/hypopnea. For example, an SPMI equal to -10 indicates that, for that patient, subband power during epochs containing apnea/hypopnea was less than subband power during non-apnea/hypopnea epochs, and that this difference was robust. In 19 of 26 patients (73%), the median of the apnea/hypopnea sample was less than the non-apnea/hypopnea sample; in 17 of 26 (65%) patients this difference was statistically significant (p < 0.05). Across patients, we computed Pearson’s correlation coefficient between the following two variates: SPMI and classifier performance (MCC; Figure 5). We found a statistically significant negative correlation (Pearson’s correlation coefficient, rho = -0.430; p = 0.0284). This association indicates that the reduction in subband power during apnea/hypopnea drives classifier performance (Figure 5). We also compared EMG subband power in epochs labeled “apnea/hypopnea” with that of epochs labelled “non-apnea/hypopnea”. Results were similar (Figure 5): In 19 of 26 patients (73%), the median of the former sample was less than the latter sample; in 14 of 26 patients (54%) this difference was statistically significant (p < 0.05); Pearson’s correlation coefficient between Wilcoxon rank sum test statistic and classifier performance gave statistically significant negative correlation (rho = -0.427; p = 0.0294). There was a clear association between EEG and EMG SPMI (Pearson’s rho = 0.978; p = 9.85e-18).

**Figure 5:**
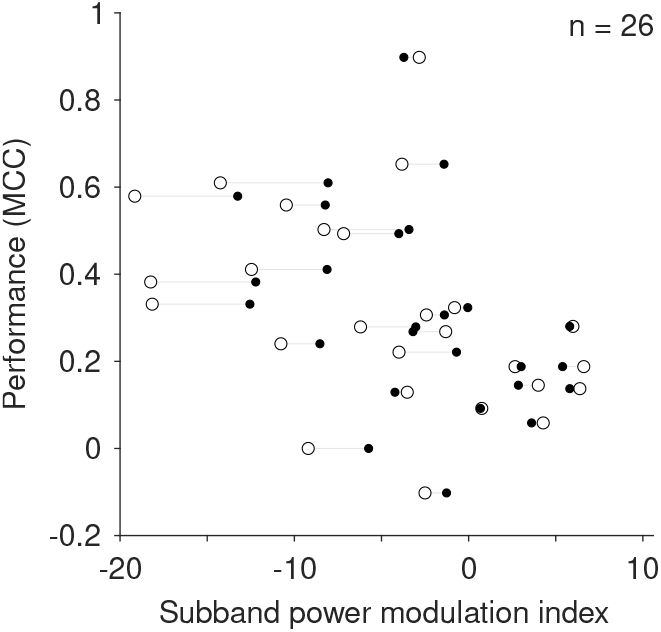
Scatter plot illustrating that classifier performance is associated with the subband power modulation index (SPMI). Each patient (n = 26) is represented by a dipole: an open circle and a closed circle joined by a line segment. Open circles represent the analysis of EEG recordings, and closed circles represent the analysis of EMG recordings. A more negative SPMI indicates a more robust difference between apnea/hypopnea versus non-apnea/hypopnea power spectra; e.g., SPMI = -10 indicates that subband power during epochs containing apnea/hypopnea was less than subband power during epochs without apnea/hypopnea, and that this difference was robust (see text). For both EEG and EMG recordings, the association between performance and SPMI is statistically significant (see text).

To better understand SA markers contained in EEG and EMG, we performed an apnea/hypopnea event-triggered average of broadband EEG and EMG power. When we used apnea/hypopnea event beginning as a trigger, we found power was relatively high prior to beginning, and relatively low thereafter. When we used apnea/hypopnea event end as a trigger, we found power was relatively low prior to event end, and relatively high thereafter (Figure 6).

**Figure 6:**
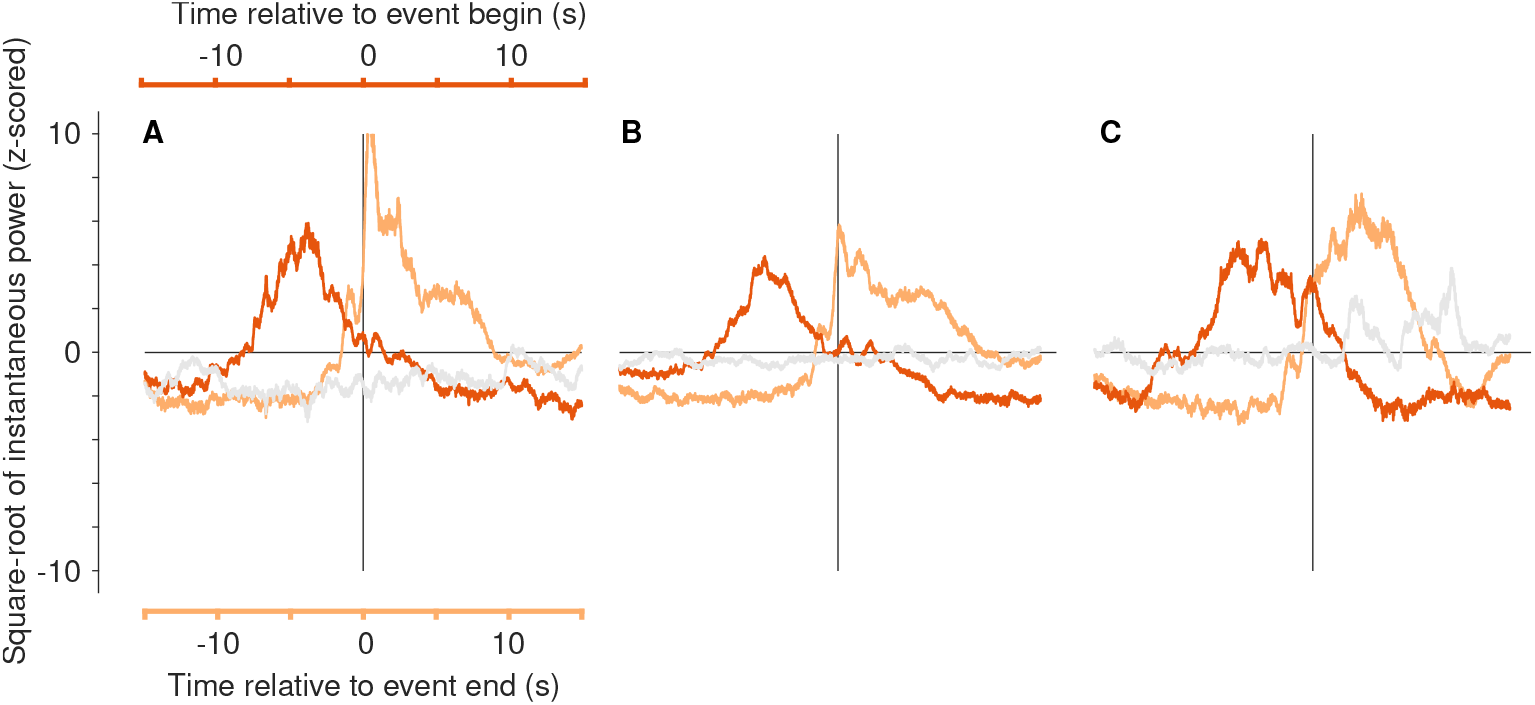
Apnea/hypopnea event-triggered average EEG (A, B) and EMG (C) broadband instantaneous power for example patient P2. (A) The dark orange trace shows power on occipital channel O1 relative to apnea/hypopnea event beginning (upper x-axis). This patient experienced 320 apnea or hypopnea events during the analysis period, and therefore this trace trace is the average of 320 individual traces. Notably, power is relatively high prior to apnea/hypopnea event beginning (time < 0 s), and relatively low thereafter (time > 0 s). The light orange trace (also an average of 320 individual traces) shows power on O1 relative to apnea/hypopnea event end (lower x-axis). Notably, power is relatively low prior to event end (time < 0 s), and relatively high thereafter (time > 0 s). The light gray trace illustrates a control analysis wherein 320 trigger times were chosen randomly from the analysis period (see Methods). By comparison to the orange traces, this gray trace shows negligible modulations of instantaneous power. (B) As in (A), however the traces are averaged across all six EEG recording channels for example patient P2. (C) As in (B), however the traces are averaged across all two EMG recording channels for example patient P2. To quantify the modulation of average EEG (B) and EMG (C) broadband instantaneous power relative to the beginning or end of apnea/hypopnea events, we computed instantaneous power modulation indices (see text).

To quantify the modulation of average EEG (Figure 6B) and EMG (Figure 6C) broadband instantaneous power relative to the beginning or end of apnea/hypopnea events, we computed instantaneous power modulation indices (IPMI). To do so, for the dark orange traces in Figure 6B and C, we summed all values occurring between time = 0 s and time = 10 s and subtracted the sum of all values occurring between time = -10 s and time = 0 s. Thus, IPMI is negative when the average instantaneous power is relatively high for t < 0 seconds, and relatively low thereafter. Similarly, this IPMI is positive when the average power is relatively low for t < 0, and relatively high thereafter. We computed IPMI in the same way for the light orange and gray traces in Figure 6B and C. For the dark-orange, light-orange, and gray traces representing EEG broadband power (Figure 6B), the modulation indices are -2.49, 4.26, and -0.24, respectively. For the dark-orange, light-orange, and gray traces representing EMG broadband power (Figure 6C), the modulation indices are -3.10, 5.76, and -0.14, respectively. This pattern of power modulation was consistent across the cohort, and is illustrated in Figure 7.

**Figure 7:**
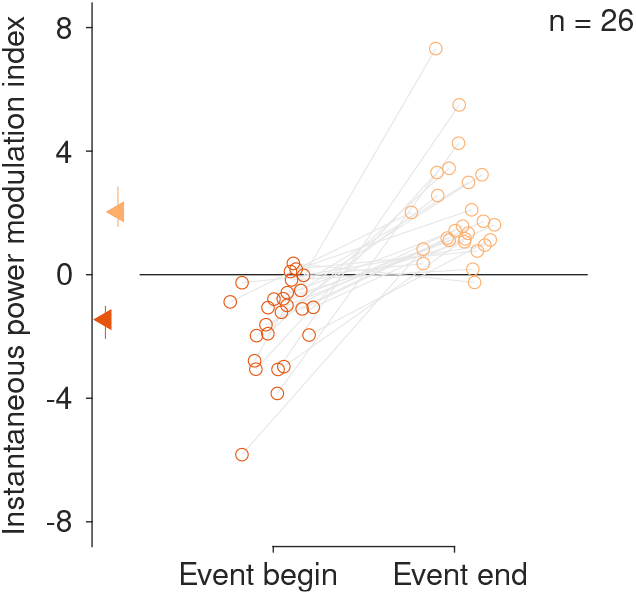
Apnea and hypopnea event beginning and end are associated with modulations of EMG broadband instanta-neous power. Each patient is represented by two symbols joined by a line segment. For each patient, the dark orange symbol (left) represents the instantaneous power modulation index computed at the beginning of apnea/hypopnea events (see Figure 6). Negative indices indicate that the event-triggered average EMG broadband instantaneous power was relatively high immediately prior to apnea/hypopnea event beginning, and relatively low thereafter (see Figure 6). At event beginning, the index is negative for most (n = 23) but not all (n = 3) patients (sign test, p = 8.7976e-05). For each patient, the light orange symbol (right) represents the index computed at the end of apnea and hypopnea events. Positive indices indicate that the event-triggered average power was relatively low immediately prior to apnea/hypopnea event end, and relatively high thereafter (see Figure 6). At event end, the index is positive for most (n = 25), but not all (n = 1), patients (sign test, p = 8.0466e-07). The triangles mark mean indices: -1.45 (apnea/hypopnea begin) and 2.03 (apnea/hypopnea end); error bars show 95% confidence intervals computed with bootstrapping (Efron and Tibshirani, 1994). The mean within-patient index difference (end minus begin) is statistically significantly different from zero (t-test, *t*_25_ = 6.70, p = 5.0478e-07).

## 4 Discussion

We have presented four models that detect apnea/hypopnea events using overnight EEG, EMG, and sleep stage labels; these models generalise to previously unseen patients. The above-chance performance of Model 3 – a simple decision strategy contingent on sleep stage label – shows that sleep stage label alone is sufficient to detect apnea/hypopnea events. Whilst EMG and EEG subband power and sample entropy can be used to detect apnea/hypopnea events (Models 1 and 2, respectively), the latter likely reflects contamination of EEG by EMG. Our reasoning is as follows:

1. EMG subband power and sample entropy can be used to detect apnea/hypopnea events at rates above chance (Figure 4, “Model 2”).
2. EEG subband power and sample entropy can be used to detect apnea/hypopnea events at rates above chance and roughly equal to that of EMG features (Figure 4, “Model 1”).
3. EEG and EMG recordings respond similarly to apnea/hypopnea events: for both signals, subband power is reduced during events (Figure 5); and for both signals, instantaneous power decreases (increases) at the time of event begin (end) (Figure 6).
4. EMG is known to contaminate EEG (Goncharova et al., 2003).
5. Therefore, EEG subband power and sample entropy likely reflect contamination of EEG by EMG.

Apnea/hypopnea event detection using EEG (or EMG) features was not correlated with detection performance using sleep stage label. Therefore, it was unsurprising that, when EEG (or EMG) features were combined with sleep stage labels, detection performance was further improved (Results and Model 4). Taking together these results, both hypotheses provided in the Introduction passed the test. This indicates that neither EEG subband power nor sample entropy are necessarily markers of SA that enable robust patient-wise detection of apnea/hypopnea events. Machine learning that uses overnight EEG for apnea/hypopnea event detection must be explainable; it must account for EMG contamination and sleep stage.

The conventional view holds that EMG, relative to EEG, is high-amplitude and high-frequency. Nonetheless, several studies have shown that EMG and EEG spectra overlap substantially, and therefore EMG cannot be eliminated from EEG by low-pass filtering. For example, Goncharova et al. (2003) demonstrated that even “weak contraction of frontalis or temporalis [facial] muscles affects spectra [of scalp recordings] across a broad frequency range that includes EEG frequency bands” (p. 1585). Specifically, those authors showed that, during facial muscle contraction, the spectra of recordings, whether from the face or the scalp, “peak at about 30 Hz and then decline steadily as frequency rises, except for a smaller peak at 45 - 70 Hz with stronger contractions. The peak frequency increases with contraction level, from 20 - 25 to 35 - 38 Hz” (p. 1583). These peak frequencies are within the range of frequencies that we analyze here (i.e., < 48 Hz; Table 1). We analysed EMG using subbands typical of EEG analyses (Table 1) to demonstrate that, in those subbands, EMG carries sufficient information to classify apnea/hypopnea at rates above chance. This demonstration adds heft to our argument: here, classification using putative EEG recordings is, in fact, classification using EMG. We note that there may exist a more judicious choice of subbands for EMG analysis, and that this more judicious choice might further improve the performance of Model 2, which used EMG subband power and sample entropy (see Methods).

Albeit here we found that overnight EMG, not EEG, subband power and sample entropy may be useful to detect apnea/hypopnea events, it is not unreasonable to suppose that apnea/hypopnea events can be detected using EEG. By analysing the power spectra of overnight EEG recordings, several groups have reported differences between cohorts of SA sufferers and normal controls, or between cohorts of sufferers of mild and severe disease (reviewed by D’Rozario et al., 2017; reviewed by Puskás et al., 2017). However, the literature is somewhat inconsistent, specifically, with regard to which frequency bands are affected by SA; whether, there, power is increased or decreased; and whether these effects are consistent between REM and NREM sleep. As noted by D’Rozario and colleagues (2017): “Of the current evidence surrounding qEEG [quantitative EEG] measures during sleep [in SA], there are substantial inconsistencies in the methodologies and a paucity of data, which makes drawing conclusions difficult.” We used a study by Jones et al. (2014) to motivate our analysis of subband power features. Specifically, Jones et al. (2014) reported a case-control study involving nine sufferers of obstructive sleep apnea. Relative to controls matched for age and body mass, sufferers’ overnight high-density EEG recorded during NREM sleep showed reduced broadband power in a circumscribed region over parietal cortex. Although our finding apnea/hypopnea events are associated with reduced EEG subband power is consistent with Jones et al. (2014), we found that these events are also associated with the reduced EMG subband power, and that EEG and EMG subband power reduction was correlated within patient. This indicates that our EEG finding is a result of EMG contamination.

In addition to analysing EEG and EMG subband power, we measured changes in EEG and EMG instantaneous power with reference to apnea/hypopnea event beginning and end. We found a robust association between instantaneous power modulations, and the timing of events of apnea/hypopnea (Figures 6 and 7). Connecting these modulations to power spectral analysis (discussed in the previous paragraph) is not straightforward; equal and opposite modulations might not be reflected in the power spectrum of a 30-second epoch. A small number of articles report EEG alterations during events of apnea/hypopnea; connecting the modulations we found to these reports is more straightforward. Walsleben et al. (1993) reported an overall decrease in EEG power during apnea/hypopnea events, consistent with our findings (Figures 6 and 7). Svanborg and Guilleminault (1996) found, during NREM apnea/hypopnea events, an increase in EEG delta-band power, on average, 13 seconds after apnea beginning. This result is potentially consistent with our findings, however, the timing of this increase with respect to apnea/hypopnea event end is unknown. Morisson et al. (1998) found, during REM apnea/hypopnea events, increased EEG delta-band power in SA sufferers versus control. This latter result is difficult to reconcile with our own (Figures 6 and 7), partly because the timing of this increase with respect to apnea/hypopnea event start and end is unknown.

The classifier presented by Zhao et al. (2021; see Introduction) outperformed ours. However, this discrepancy is not surprising for several reasons. First, we used leave-one-patient-out cross-validation (“patient-wise cross-validation”). This approach provides a better measure of the external validity of a classification model, that is, whether the model is likely to generalise to previously unseen patients and, in turn, be useful in a clinical setting. By contrast, Zhao and colleagues pooled data across patients before performing cross-validation. Performance is the price paid for external validity. Second, our patient cohort was consecutive — patients reported here were analyzed in the order they were provided to the principal investigator (LH) by sleep physiologists who were naive to the study’s hypotheses — whereas Zhao et al. did not analyze patients consecutively. Performance, also, is the price paid to mitigate selection bias. We emphasize that, here, our aim was not to maximize classifier performance — it is likely that there exist other informative features (beyond subband power and sample entropy) to aid performance, and although a RF classifier has been shown to outperform several other types of classifier (Zhao et al., 2021), a RF classifier is unlikely to be optimal. Rather, we aimed to better understand the putative EEG, and actual EMG, signals enabling patient-wise detection of epochs containing apnea/hypopnea events, and to partition the contribution of sleep.

## 5 Conclusion

Previous work using overnight EEG recordings to detect which 30-second epochs contain apnea/hypopnea events has lacked mechanistic explanation. This is a problem because it is unclear whether there is, in fact, a robust EEG marker of apnea/hypopnea events which enables detection/classification that generalises to new patients; specifically, it is unclear whether EEG subband power and sample entropy are robust markers. Sleep stage and EMG are reflected in EEG recordings, and we have shown here that either sleep stage or EMG (specifically, EMG subband power and sample entropy) enable detection/classification of apnea/hypopnea events that generalises to new patients. These results indicate that EEG subband power and sample entropy may not be markers of SA enabling robust apnea/hypopnea event classification, but EMG subband power and sample entropy probably are. In medicine, machine learning must be explainable (Kundu, 2021). A mechanistic understanding of the drivers of classification can be leveraged to develop hypothesis-driven classifiers; future work using overnight EEG to detect SA must specifically account for sleep stage and EMG contamination of EEG recordings.

## Data Availability

Data produced in the present study are available upon reasonable request to corresponding author.

## Author contributions

LH and AV were involved in the conception and the design of the project; LH, AV, DZ and JZ were involved in the project’s execution; LH, DZ, JZ and JW were involved in the data analysis; LH, AV, DZ and JZ were involved in the interpretation of data; all authors were involved in drafting, writing or revising the research report, and read and approved the final version of the manuscript.

